# Effectiveness of successive booster vaccine doses against SARS-CoV-2 related mortality in residents of Long-Term Care Facilities in the VIVALDI study

**DOI:** 10.1101/2023.03.01.23286627

**Authors:** Oliver Stirrup, Madhumita Shrotri, Natalie L. Adams, Maria Krutikov, Borscha Azmi, Igor Monakhov, Gokhan Tut, Paul Moss, Andrew Hayward, Andrew Copas, Laura Shallcross

## Abstract

We evaluated the effectiveness of 1-3 booster vaccinations against SARS-CoV-2 related mortality among a cohort of 13407 older residents of long-term care facilities (LTCFs) participating in the VIVALDI study in England in 2022. Cox regression was used to estimate relative hazards of SARS-CoV-2 related death following booster vaccination relative to 2 doses (after 84+ days), stratified by previous SARS-CoV-2 infection and adjusting for age, sex and LTCF capacity. Each booster provided additional short-term protection relative to primary vaccination, with consistent pattern of waning to 45-75% reduction in risk beyond 112 days.

## Introduction

Long-term care facilities (LTCFs) in the UK were severely impacted by COVID-19 early in the pandemic[1]. As such, LTCF staff and residents were prioritised for primary vaccination against SARS-CoV-2 starting in December 2020[2] and for additional rounds of booster vaccination from September 2021, March 2022 and September 2022.

We previously reported waning of protection against infection and severe clinical outcomes following primary course vaccination in LTCF residents from 84 days (12 weeks) following second dose [3], with an improvement in protection against severe outcomes observed following first booster vaccine dose (third dose)[3] that remained after emergence of the Omicron variant[4]. This study aims to produce an updated evaluation of the effectiveness of third, fourth and fifth dose booster vaccination against SARS-CoV-2 associated death amongst residents of LTCFs in England over the year 2022.

We focused on the outcome of mortality because of changes to testing policy in LTCFs in the UK within the period analysed, which make it challenging to analyse effects on SARS-CoV-2 infection incidence or to evaluate hospital admissions in a timely manner.

## Methods

VIVALDI is a prospective cohort study investigating SARS-CoV-2 in LTCF residents and staff in England [5]. For this study, the analysis period is defined from January 1 2022 to 31 December 2022, and residents aged >65 years were eligible for inclusion if they had at least one polymerase chain reaction (PCR) or lateral flow device (LFD) result linked to a participating LTCF available within this period. Following national guidelines, residents underwent monthly routine PCR testing until the end of March 2022, when the policy switched to symptomatic and outbreak testing only. The analysis period was chosen to allow our prior estimates of vaccine effectiveness to be updated whilst retaining a period when asymptomatic testing was still in use, allowing identification of the cohort of residents in participating LTCFs in early 2022. As previously, we excluded individuals who had not received at least two vaccine doses before the analysis period[4] because unvaccinated residents are substantially more likely to be receiving end of life care.

As previously we retrieved all available PCR and LFD results from the national testing programme through the COVID-19 Datastore. Test results and vaccination (National Immunisation Management Service) and mortality (Office for National Statistics) data from national records were linked to study participants using pseudo-identifiers based on National Health Service (NHS) numbers[3]. COVID-19 death was defined as death within 28 days of positive PCR or LFD test or with COVID-19 recorded as primary or secondary cause of death on the death certificate. SARS-CoV2 serological test results for IgG antibodies to the nucleocapsid protein (Abbott ARCHITECT system (Abbott, Maidenhead, UK)) were linked in a subset of participants who consented to blood sampling specifically for the VIVALDI study[6].

### Statistical analysis

We examined individual-level vaccine effectiveness against SARS-CoV-2 associated death. Individuals were eligible for inclusion if they had complete data on sex and age, had received at least 2 vaccine-doses 84 days before analysis start date, had at least one PCR or LFD test result recorded within the analysis period and were linked to a LTCF participating in VIVALDI with data on total number of staff and residents. Individuals with third vaccine dose recorded before September 14 2021, fourth dose before March 21 2022 and fifth dose before September 5 2022 were excluded as these dates corresponded to national roll-out to residents of these doses.

We used Cox regression models to derive adjusted hazard ratios (HRs) for risk of SARS-CoV-2 linked death. Vaccination status was included as a time-varying covariable. The reference category was 2 vaccine doses, with ≥84 days elapsed from Dose 2 (although this was varied to create graphical summaries of the estimated effect of successive doses). Exposure categories were 0–13, 14–48, 49–83, 84-111, 112-139 and ≥140 days following Dose 3 and Dose 4, and 0–13, 14–48, 49–83 and ≥84 days following Dose 5. Individuals entered the risk period on 1 January 2022, or date of their first recorded PCR/LFD result within VIVALDI if later. Individuals with positive PCR/LFD result within 30 days prior to 1 January 2022 entered the risk period from the 31st day post-positive test.

Individuals exited the risk period at the earliest of: death or end of analysis period. Individuals were additionally censored at 29 days post-positive SARS-CoV-2 test if they survived to this point; COVID-related death from the original infection would be possible beyond this time but the individual would also be at lower risk for a new SARS-CoV-2 infection, and so censoring at 29 days was chosen to match our mortality outcome definition. Baseline hazard was defined over calendar time. 95% CIs were calculated using robust SEs accounting for dependence of infection events within LTCFs. Analysis was stratified by evidence of SARS-CoV2 infection prior to the risk period. We adjusted for sex (binary variable), age (5-knot restricted cubic spline term) and LTCF size expressed as total number of beds (linear term). All statistical analyses were conducted using STATA 17.0.

### Patient Consent Statement

Patient consent was not obtained for data use other than for those patients who underwent blood sampling specifically for the VIVALDI study. The legal basis to access data from residents without informed consent is provided by Health Research Authority Confidentiality Advisory Group approval (21/CAG/0156). Ethical approval was obtained from South Central-Hampshire B Research Ethics Committee (20/SC/0238).

## Results

13407 residents from 327 LTCFs were included, following exclusion of 1051 because 2-dose vaccination had not been completed more than 84 days before start of the analysis period, 194 because booster vaccination doses were recorded prior to national roll-out and 904 for whom a positive SARS-CoV-2 test was recorded on the first day of their at-risk period. 3411 residents (25.4%) had recorded evidence of SARS-CoV-2 infection prior to the analysis period. The median age was 86.6 years (IQR, 80.2-91.8). 4132 residents died during the analysis period, of which 428 were associated with SARS-CoV-2.

12072 (90.0%) residents had received third-dose booster vaccination dose prior to the analysis period, and 13104 (97.7%) by the end (98.8% of those surviving to year-end) (Figure 1). First booster doses were Pfizer in the majority (n=12474, 95.2%), with Moderna also used (n=593, 4.5%). Second boosters (fourth dose) had been received by 10846 (80.9%, 93.0% of survivors) and third boosters (fifth dose) by 7311 (54.5%, 70.8% of survivors) of residents by end of analysis period. The majority were Moderna vaccines for second booster (6421 (59.2%)) and for third booster (5991 (81.9%)). 8621 (64.3%) residents had received a bivalent vaccine by end of study period, mostly as third booster dose but in some as first (n=120) or second (n=1455) booster received.

**Figure 1.**
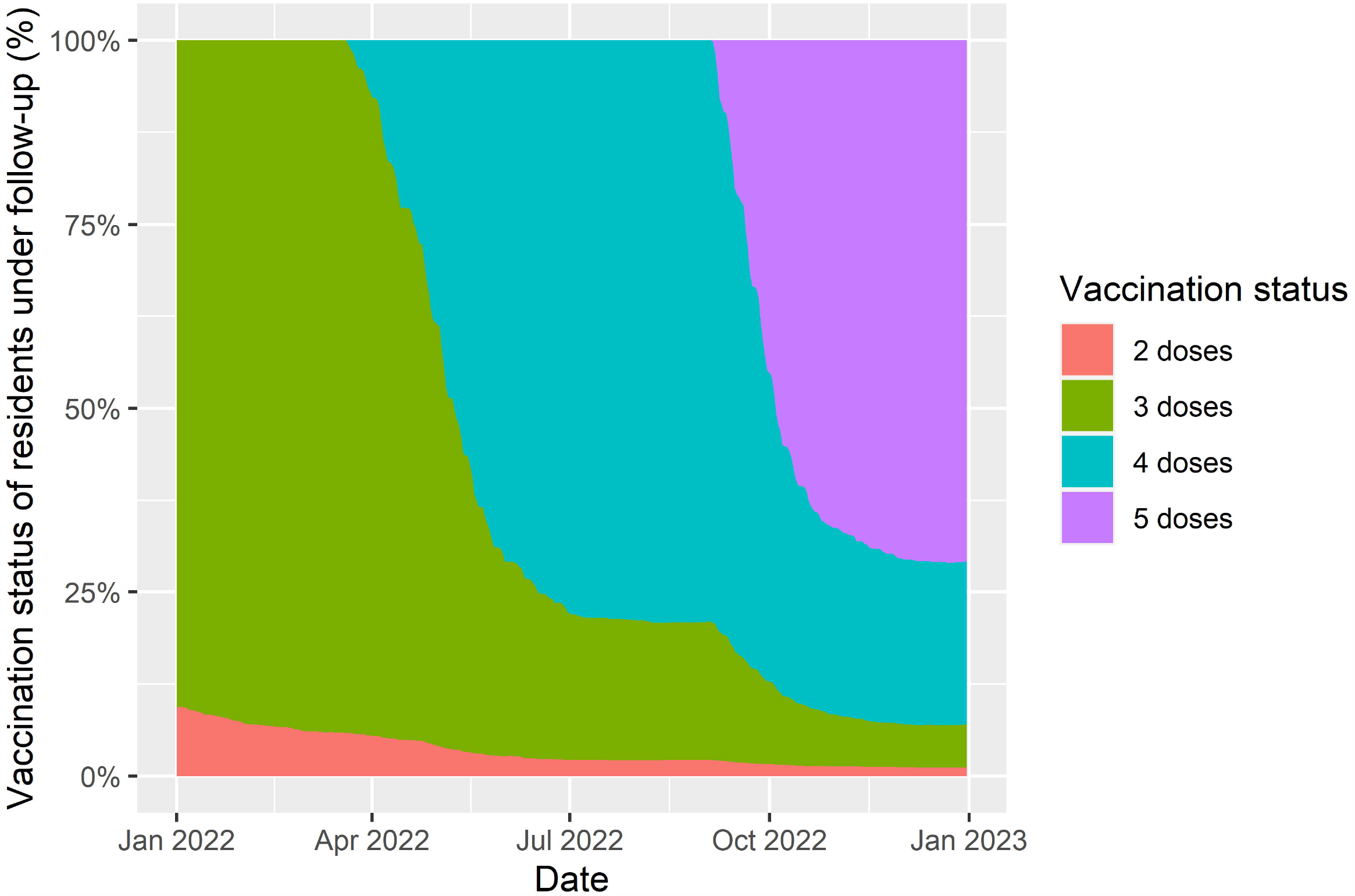
Plot of booster vaccination status of residents of long term care facilities included in the analysis. The percentage of residents with each number of vaccine doses is given among those in follow-up for the analysis at any given point in time.

In residents without known prior SARS-CoV-2 infection, first booster reduced risk of SARS-CoV-2 linked death after 0-13 days (none observed), 14-48 days (HR 0.20, 0.07-0.58), 49-83 days (0.25, 0.13-0.47) and 84-111 days (0.30, 0.18-0.51), with some apparent waning in level of protection by 112-139 days (0.44, 0.28-0.69) and 140+ days (0.38, 0.24-0.61) (Figure 2, Table 1). A similar pattern was observed following fourth and fifth dose vaccination, although confidence intervals for HRs were wider for fourth dose beyond 84 days and for fifth dose (Figure 2d) because of lower available follow-up time and lower incidence of SARS-CoV-2 infection in the latter half of 2022. Residents with known infection prior to the analysis period were at reduced risk of death relative to those without prior infection (0.55,0.28-1.08; among those with 2-dose vaccination). Within this group, the pattern of further protection from booster vaccination was similar to those without known prior infection, although there is greater uncertainty in estimates. Adding a parameter to the model representing receipt of a bivalent (BA.1) vaccine indicated possible greater protection but with a wide confidence interval (0.81, 0.29-2.25).

**Table 1.**
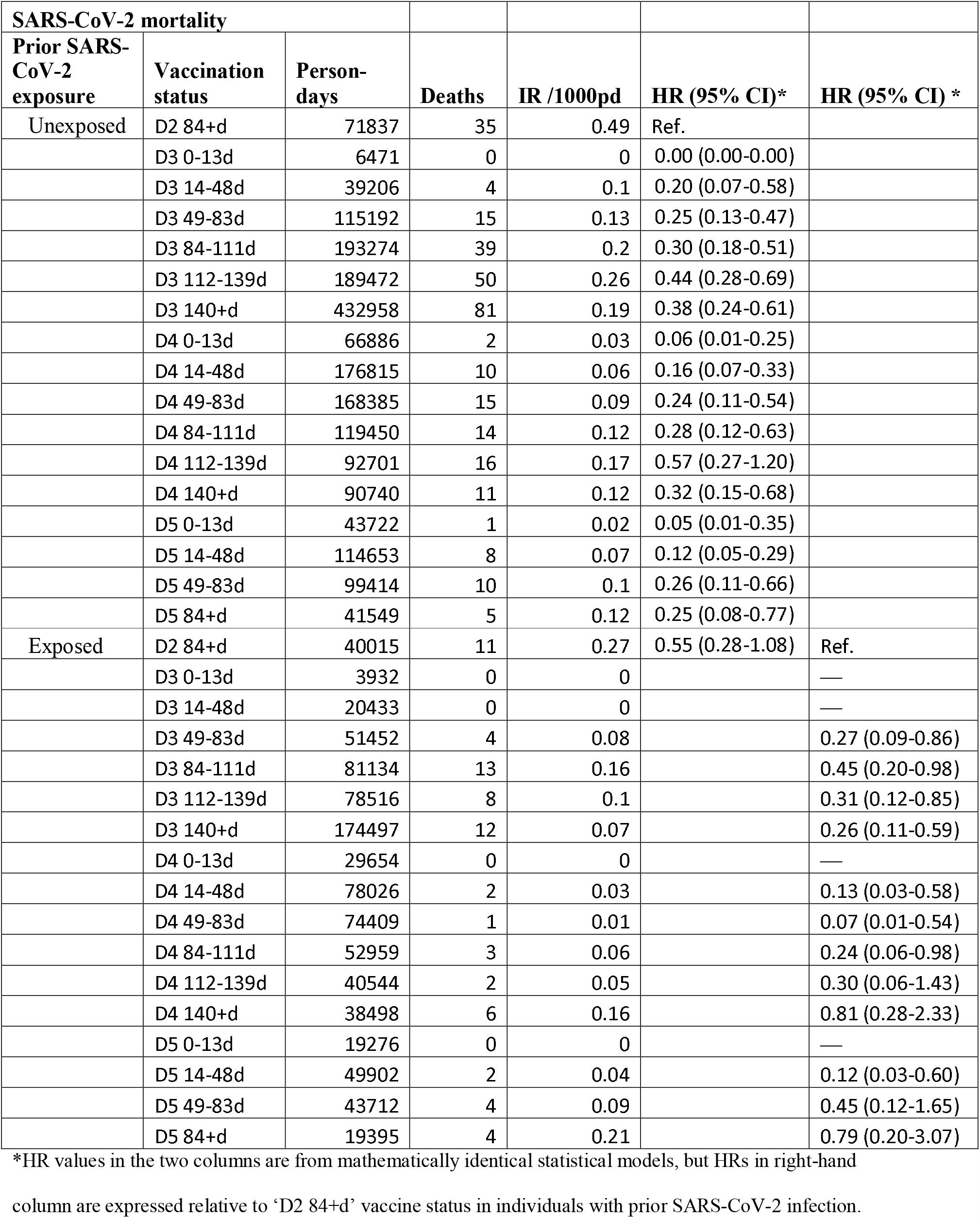
Crude event rates and adjusted hazard ratios against SARS-CoV-2 associated death (within 28 days of a positive PCR or LFD test, and/or recorded on death certificate) for LTCF residents, by prior SARS-CoV-2 exposure, and vaccination status

| SARS-CoV-2 mortality |  |  |  |  |  |  |
| --- | --- | --- | --- | --- | --- | --- |
| Prior SARS-CoV-2 exposure | Vaccination status | Person-days | Deaths | IR /1000pd | HR (95% CI)* | HR (95% CI) * |
| Unexposed | D2 84+d | 71837 | 35 | 0.49 | Ref. |  |
|  | D3 0-13d | 6471 | 0 | 0 | 0.00 (0.00-0.00) |  |
|  | D3 14-48d | 39206 | 4 | 0.1 | 0.20 (0.07-0.58) |  |
|  | D3 49-83d | 115192 | 15 | 0.13 | 0.25 (0.13-0.47) |  |
|  | D3 84-111d | 193274 | 39 | 0.2 | 0.30 (0.18-0.51) |  |
|  | D3 112-139d | 189472 | 50 | 0.26 | 0.44 (0.28-0.69) |  |
|  | D3 140+d | 432958 | 81 | 0.19 | 0.38 (0.24-0.61) |  |
|  | D4 0-13d | 66886 | 2 | 0.03 | 0.06 (0.01-0.25) |  |
|  | D4 14-48d | 176815 | 10 | 0.06 | 0.16 (0.07-0.33) |  |
|  | D4 49-83d | 168385 | 15 | 0.09 | 0.24 (0.11-0.54) |  |
|  | D4 84-111d | 119450 | 14 | 0.12 | 0.28 (0.12-0.63) |  |
|  | D4 112-139d | 92701 | 16 | 0.17 | 0.57 (0.27-1.20) |  |
|  | D4 140+d | 90740 | 11 | 0.12 | 0.32 (0.15-0.68) |  |
|  | D5 0-13d | 43722 | 1 | 0.02 | 0.05 (0.01-0.35) |  |
|  | D5 14-48d | 114653 | 8 | 0.07 | 0.12 (0.05-0.29) |  |
|  | D5 49-83d | 99414 | 10 | 0.1 | 0.26 (0.11-0.66) |  |
|  | D5 84+d | 41549 | 5 | 0.12 | 0.25 (0.08-0.77) |  |
| Exposed | D2 84+d | 40015 | 11 | 0.27 | 0.55 (0.28-1.08) | Ref. |
|  | D3 0-13d | 3932 | 0 | 0 |  | — |
|  | D3 14-48d | 20433 | 0 | 0 |  | — |
|  | D3 49-83d | 51452 | 4 | 0.08 |  | 0.27 (0.09-0.86) |
|  | D3 84-111d | 81134 | 13 | 0.16 |  | 0.45 (0.20-0.98) |
|  | D3 112-139d | 78516 | 8 | 0.1 |  | 0.31 (0.12-0.85) |
|  | D3 140+d | 174497 | 12 | 0.07 |  | 0.26 (0.11-0.59) |
|  | D4 0-13d | 29654 | 0 | 0 |  | — |
|  | D4 14-48d | 78026 | 2 | 0.03 |  | 0.13 (0.03-0.58) |
|  | D4 49-83d | 74409 | 1 | 0.01 |  | 0.07 (0.01-0.54) |
|  | D4 84-111d | 52959 | 3 | 0.06 |  | 0.24 (0.06-0.98) |
|  | D4 112-139d | 40544 | 2 | 0.05 |  | 0.30 (0.06-1.43) |
|  | D4 140+d | 38498 | 6 | 0.16 |  | 0.81 (0.28-2.33) |
|  | D5 0-13d | 19276 | 0 | 0 |  | — |
|  | D5 14-48d | 49902 | 2 | 0.04 |  | 0.12 (0.03-0.60) |
|  | D5 49-83d | 43712 | 4 | 0.09 |  | 0.45 (0.12-1.65) |
|  | D5 84+d | 19395 | 4 | 0.21 |  | 0.79 (0.20-3.07) |

\*HR values in the two columns are from mathematically identical statistical models, but HRs in right-hand
column are expressed relative to 'D2 84+d' vaccine status in individuals with prior SARS-CoV-2 infection.

**Figure 2.**
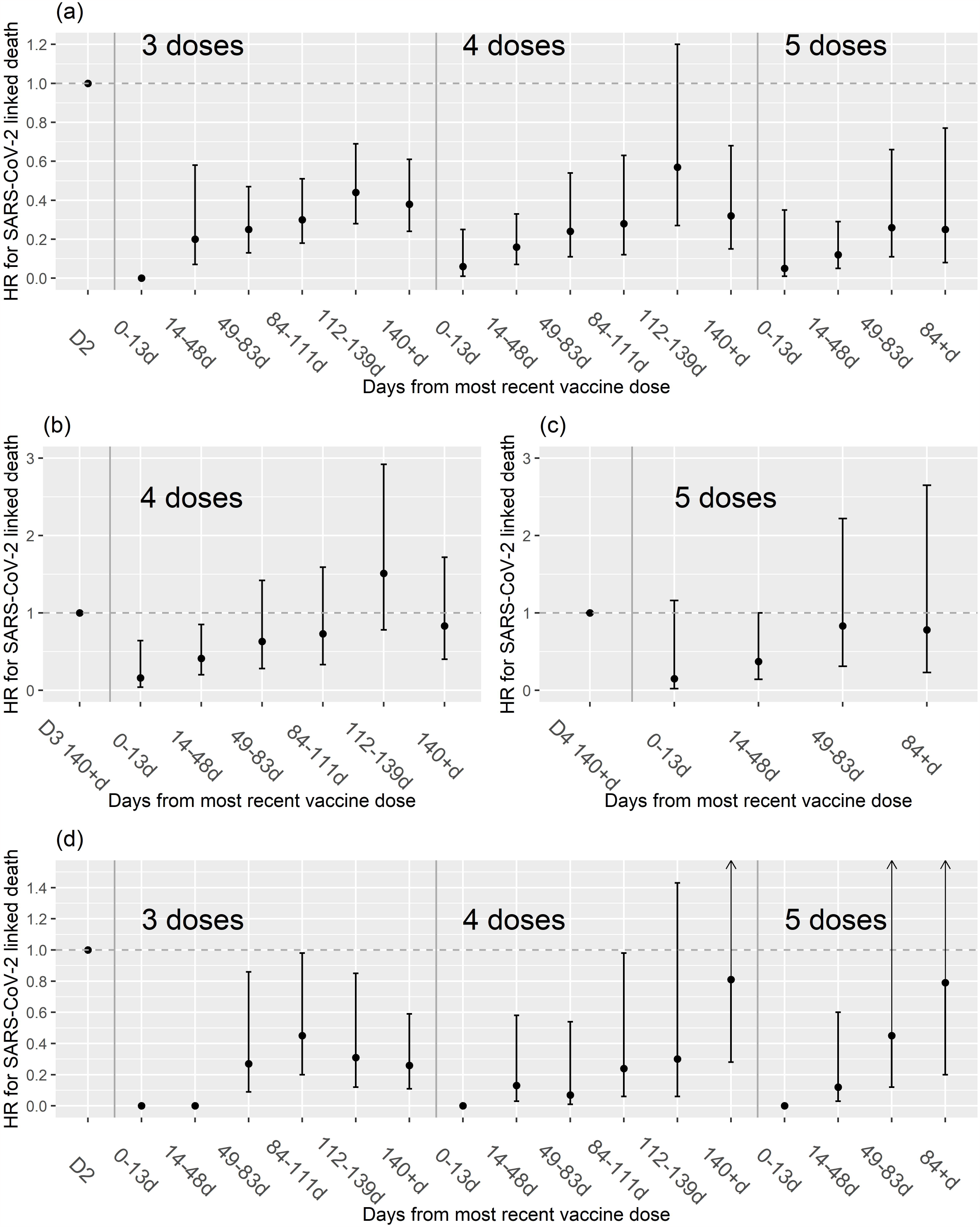
Plot of estimated hazard ratio (HR) for SARS-CoV-2 linked death (within 28 days of a positive PCR or LFD test, and/or recorded on death certificate) following receipt of booster vaccine doses, among long term care facility residents without evidence of prior SARS-CoV-2 infection (a-c) and in those with prior SARS-CoV-2 infection (d). Results are shown for 3-5 doses relative to 2-dose vaccination in (a) and (d), for 4 doses relative to 3 doses beyond 140 days in (b) and for 5 doses relative to 4 doses beyond 140 days in (c).

## Discussion

We found evidence that third, fourth and fifth dose booster vaccination provide additional short-term protection against SARS-CoV-2 linked mortality among LTCF residents, relative to primary vaccination only. This is consistent with data on fourth dose vaccination of LTCF residents in the USA[7] and Canada[8]. The pattern of waning of protection appeared to be similar for successive booster doses, stabilising at a 45-75% reduction in risk beyond 112 days (16 weeks).

Our data suggest that the combination of booster vaccination with prior SARS-CoV-2 infection provides particularly strong protection against subsequent mortality from the virus, in line with previous studies in the general population[9]. We are likely to have underestimated the protective effect of prior infection in this analysis, as we have previously found the cumulative incidence of detected SARS-CoV-2 infection to be substantially higher among residents who have undergone testing for anti-nucleocapsid antibodies[6]. This suggests that many of the ‘unexposed’ group in our analyses may have in fact been infected prior to the risk period, although the prevalence of prior infection may be lower in residents who were admitted to care homes in the second half of 2021 and 2022 compared to those resident since 2020.

It is possible that the primarily bivalent fifth dose vaccination provided some additional long-term protection, but the pattern of reduction in risk over time did not differ markedly from that observed for prior booster doses. This is consistent with data from the general population in the USA[10], perhaps due to the fact that different Omicron sub-lineages were circulating by the time that bivalent vaccines based on the BA.1 lineage were rolled out[11].

## Supporting information

STROBE checklist

## Data Availability

De-identified test results and limited meta-data will be made available for use by researchers in future studies, subject to appropriate research ethical approvals, once the VIVALDI study cohort has been finalised.

## Acknowledgements

We thank the staff and residents in the long-term care facilities who participated in this study and Mark Marshall at National Health Service (NHS) England who pseudonymised the electronic health records. The views expressed in this publication are those of the authors and not necessarily those of the NHS or the UK Health Security Agency.

